# Quantifying the impact of real-world evidence: the sacubitril/valsartan experience

**DOI:** 10.1101/2024.09.27.24314305

**Authors:** Andrew J. Epstein, Morgan H. Persky, Saif S. Rathore

## Abstract

**Background:** The impact of RWE publication on use of approved therapies—including the existence of an effect and timing of impact—is unknown.

**Methods:** We assessed the relationship between the cadence of RWE publications and sales of sacubitril/valsartan (Entresto®) following launch. Publication data were derived from a PubMed search over 7/01/2015–12/31/2023 for English-language, human subjects’ studies with the drug name (Entresto® or sacubitril) and indication (heart failure) in the title or abstract. Quarterly RWE publication data were combined with quarterly information on sacubitril/valsartan sales over the 34-quarter period spanning Q3 2015 through Q4 2023. Quarter-to-quarter change in total sales was modeled with linear regression as a function of counts of previously published RWE-related studies, adjusting for a linear time trend to control for any underlying trends in sales growth; change in total sales from the quarter prior to publication to the quarter of publication to control for recent sales changes; and the cumulative count of RWE publications from Q3 2015 through the previous quarter to control for the contribution of previous RWE publications to future sales.

**Results:** The PubMed search identified 888 studies; of these, 333 (37.5%) were classified as RWE-related. An additional RWE-related publication (in quarter q) was associated with an estimated mean increase in quarterly sales of $1.8M (from quarter q to q+1 following publication), $3.5M (from q+1 to q+2), and $2.7M (from q+2 to q+3). In adjusted models, an additional RWE publication was associated with a mean gain in sales from quarter q+1 to q+2 of $2.6M (95% CI −$0.2M to $5.4M). RWE publications were not associated with an increase in sales in the quarter following publication, and the association was attenuated by third quarter following publication.

**Conclusion:** RWE studies were associated with increased use of sacubitril/valsaratan, providing preliminary evidence of RWE’s ability to support therapy adoption.

## Introduction

Real-world evidence (RWE) can demonstrate safety, effectiveness, and value following a new therapy’s approval and launch.^1^ The FDA has prioritized RWE programs to regulate approved therapies and products and has sometimes expanded approved indications based on RWE.^2^ Recognizing RWE’s distinct value, manufacturers, payers, and providers have invested ∼$18B in RWE in 2024, with growth forecasted to increase 13% annually through 2032.^3^ Although publications of clinical trial results have been linked to drug sales,^4^ the influence of RWE on the use of approved therapies is unknown. This is especially important for therapies with novel mechanisms of action in competitive therapeutic spaces.

We sought to assess the relationship between the cadence of RWE publications and subsequent sales of sacubitril/valsartan (Entresto®). As a first-in-class angiotensin receptor-neprilysin inhibitor approved for heart failure treatment, sacubitril/valsartan entered a therapeutic area with multiple established therapies. We evaluated overall drug sales following peer-reviewed RWE publications, including timing to and gain in sales following publication.

## Methods

Publication data were derived from a PubMed search over 7/01/2015–12/31/2023 for English-language, human subjects’ studies with the drug name (Entresto® or sacubitril) and indication (heart failure) in the title or abstract. Studies without an abstract were excluded. Studies were classified as RWE-related if the abstract had at least one keyword from this list: “clinical,” patient,” “treatment,” “hospital,” “physician,” “visit,” and “admission,” and at least one keyword from this list: “real world,” “real-world,” “observation,” “cohort,” “practice,” “effectiveness,” “registry,” and “cost-effective.” Candidate abstracts were reviewed by an author (SSR) to confirm that they contained RWE as defined by the Food and Drug Administration: “clinical evidence regarding a medical product’s use and potential benefits or risks derived from analysis of real-world data.”^5^

Quarterly RWE publication data were combined with quarterly information on sacubitril/valsartan sales over the 34-quarter period spanning Q3 2015 through Q4 2023. RWE publications were assigned to a calendar quarter based on their publication date and counted by quarter. Total quarterly sales were collected from Novartis’s US Securities and Exchange Commission 6-K quarterly reports^6^ and inflated to 2023 USD using the US Bureau of Labor Statistics’ Producer Price Index for pharmaceutical manufacturing.^7^

We estimated linear regression models of the quarter-to-quarter change in total sales as predicted by counts of previously published RWE-related studies. As it is unclear when RWE publications may influence sales (if at all), we examined unadjusted and adjusted associations between publication count in quarter q and three consecutive quarterly changes in future sales (q and q+1, q+1 and q+2, and q+2 and q+3 in separate models). To mitigate concerns of spurious correlation between simultaneously increasing trends in publications and sales, the adjusted models accounted for a linear time trend to control for any underlying trend in sales growth; the change in total sales from the quarter prior to publication to the quarter of publication (q−1 to q) to control for recent sales changes; and the cumulative count of RWE publications from Q3 2015 through the previous quarter (q−1) to control for the contribution of previous RWE publications to future sales. Standard errors were made robust to heteroskedasticity and autocorrelation for quarters q−1 through q+3.^8^

## Results

The PubMed search identified 888 studies; of these, 333 (37.5%) were classified as RWE-related. Mean quarterly publication count was 9.8 and ranged from 1 (Q3 2015) to 26 (Q3 2021).

Total quarterly sales increased steadily from $0 in Q3 2015 to $1.635B in Q4 2023; mean quarterly sales were $626M. The mean quarter-to-quarter change in sales was $50M and ranged from −$35M (Q3 2023) to $156M (Q4 2022).

In the unadjusted models, an additional RWE-related publication was associated with an estimated mean increase in quarterly sales of $1.8M (from quarter q to q+1 following publication), $3.5M (from q+1 to q+2), and $2.7M (from q+2 to q+3).

In adjusted models accounting for a secular trend in sales, the number of earlier RWE publications, and change in sales in the prior quarter, an additional RWE publication was associated with a mean gain in sales from quarter q+1 to q+2 of $2.6M (95% CI −$0.2M to $5.4M). RWE publications were not associated with an increase in sales in the quarter following publication, and the association was attenuated by third quarter following publication (**Table 1**).

**Table 1.** Impact of RWE publication and subsequent change in sacubitril/valsartan sales.

|  | Change in sales from<br>quarters q to q+1 |  | Change in sales from<br>quarters q+1 to q+2 |  | Change in sales from<br>quarters q+2 to q+3 |  |
| --- | --- | --- | --- | --- | --- | --- |
|  | Unadjusted | Adjusted | Unadjusted | Adjusted | Unadjusted | Adjusted |
| **Number of clinical pubs in quarter q** |  |  |  |  |  |  |
| Coefficient | 1.8 | −2.5 | 3.5 | 2.6 | 2.7 | 0.7 |
| 95% confidence interval | [−0.5, 4.1] | [−4.8, −0.2] | [2.4, 4.7] | [−0.2, 5.4] | [−0.1, 5.5] | [−4.1, 5.4] |
| Sample size (quarters) | 32 | 32 | 31 | 31 | 30 | 30 |

Notes:
Outcome is measured in 2023 US dollars.
Adjusted models control for a linear time trend, the change in sales from quarters q−1 to q, and the cumulative count of RWE publications through quarter q−1.
Linear regression models used Newey-West heteroskedasticity and autocorrelation consistent standard errors (up to lag 4).

## Discussion

In the 8 years following approval of sacubitril/valsartan, publication of RWE studies was associated with sales growth 2 quarters later, with an extra publication associated with an extra $2.6M in revenue after accounting for other factors. While the impact likely varies across publications, our study suggests that, on average, an additional RWE publication nominally contributes to overall therapy use. In response to documented challenges achieving goals of guideline-directed medical therapy in heart failure,^9^ our study suggests continued RWE generation may be helpful in increasing use of newer therapies.

Our study has limitations. While our study cannot demonstrate a true causal relationship, we believe it does provide suggestive evidence to support the value of RWE generation. These findings are limited to a single therapy and may not generalize to other products. Our analysis was constructed to assess the average impact across a series of publications and does not quantify the total impact of a publication on lifetime sales, which would require more data and complex analysis. Moreover, our data do not disentangle how RWE publication leads to more sales, including through any professional or commercial efforts that might have contributed to medication use.

Publication of RWE studies was associated with more use of sacubitril/valsaratan following its introduction. As such, this analysis provides novel preliminary evidence of RWE’s ability to support therapy sales and thus adoption.

## Data Availability

All data produced in the present study are available upon reasonable request to the authors.

